# Changing travel patterns in China during the early stages of the COVID-19 pandemic

**DOI:** 10.1101/2020.05.14.20101824

**Authors:** Hamish Gibbs, Yang Liu, Carl AB Pearson, Christopher I Jarvis, Chris Grundy, Billy J Quilty, Charlie Diamond, LSHTM CMMID COVID-19 working group, Rosalind M Eggo

**Author notes:** **Materials & Correspondence:** Hamish Gibbs, Yang Liu. authors contributed equally (Hamish Gibbs, Yang Liu). The LSHTM CMMID COVID-19 working group is (order determined at random): David Simons, Amy Gimma, Quentin J Leclerc, Megan Auzenbergs, Rachel Lowe, Kathleen O’Reilly, Matthew Quaife, Joel Hellewell, Gwenan M Knight, Thibaut Jombart, Petra Klepac, Simon R Procter, Arminder K Deol, Eleanor M Rees, Stefan Flasche, Adam J Kucharski, Sam Abbott, Fiona Yueqian Sun, Akira Endo, Graham Medley, James D Munday, Sophie R Meakin, Nikos I Bosse, W John Edmunds, Nicholas G. Davies, Kiesha Prem, Stéphane Hué, C Julian Villabona-Arenas, Emily S Nightingale, Rein M G J Houben, Anna M Foss, Damien C Tully, Jon C Emery, Kevin van Zandvoort, Katherine E. Atkins, Alicia Rosello, Sebastian Funk, Mark Jit, Samuel Clifford, Timothy W Russell.

## Abstract

Understanding changes in human mobility in the early stages of the COVID-19 pandemic is crucial for assessing the impacts of travel restrictions designed to reduce disease spread. Here, relying on data from mainland China, we investigated the spatio-temporal characteristics of human mobility between 1st January and 1st March 2020 and discussed their public health implications. An outbound travel surge from Wuhan before travel restrictions were implemented was also observed across China due to the Lunar New Year, indicating that holiday travel may have played a larger role in mobility changes compared to impending travel restrictions. Holiday travel also shifted healthcare pressure related to COVID-19 towards locations with lower access to care. Network analyses showed no sign of major changes in the transportation network after Lunar New Year. Changes observed were temporary and have not yet led to structural reorganisation of the transportation network at the time of this study.

**One sentence summary:** Understanding travel before, during, and after the introduction of travel restrictions in China in response to the COVID-19 Pandemic.

## Introduction

The COVID-19 pandemic began in Wuhan, China, in late 2019, came to prominence in January 2020, and quickly spread within the country. January is also a major holiday period in China, and the 40-day period around Lunar New Year (LNY), or *Chunyun*, marks the largest annual human migration in the world, with major travel flows out of large cities^1^. In 2019, nearly 3 billion individual journeys were made during *Chunyun*^2^. In 2020, *Chunyun* lasted from 10th January to 18th February^3^, with the first day of the LNY holidays on 24th January, followed by the first day of LNY on 25th January. This period coincided with the initial phase of the COVID-19 pandemic, and there has been speculation that holiday travel may have accelerated the propagation of COVID-19 both within China and internationally^4^.

As part of initial efforts to contain the outbreak, the Chinese government announced a *cordon sanitaire* for the city of Wuhan, Hubei Province, starting on 23rd January 2020, one day before LNY holidays. This intervention restricted all non-essential movement into and out of the city. Services at airports, train stations, long-distance bus stations, and commercial ports were all suspended^5^. Several studies have focused on assessing the effectiveness of the *cordon sanitaire* in Wuhan and other domestic travel restrictions in China in the context of COVID-19 control^6–8^. As other affected regions worldwide begin implementing similar travel restrictions^9^, it is critical to understand human mobility patterns during the initial phase of the COVID-19 pandemic and their potential implications for other countries.

Out-going traffic from Wuhan was reduced by 89% within two days of the *cordon sanitaire*, according to data from Baidu Huiyan, an internet service company in China which uses location targeting to provide services to users. Baidu’s Location Based Service (LBS)^10^ provides travel fluxes between prefectures in China during the annual *Chunyun* period to allow monitoring of movement of people using their services.

We used daily prefecture-level movement data across China provided by Baidu Huiyan^10^ to understand the spatial and temporal characteristics of movement patterns before, during and after the COVID-19 epidemic in Wuhan. Relying on a range of techniques from trajectory clustering to network analysis, we examined human movement on multiple geographic scales to provide a complete picture of the overall dynamics while drawing links to their public health implications.

## Results

### Human movement surrounding the epicentre – Wuhan, Hubei

Before the *cordon sanitaire* and during the initial phase of the COVID-19 epidemic, outbound travel from Wuhan was marked by an early-January peak, followed by a sharper second peak in the days before the LNY holidays (Figure 1a). The first peak was not observed in 2019, while the second peak was higher in 2020 than 2019. Because the start of Wuhan’s *cordon sanitaire* and the beginning of LNY holidays were only one day apart, we refer only to LNY while describing our results.

**Figure 1.**
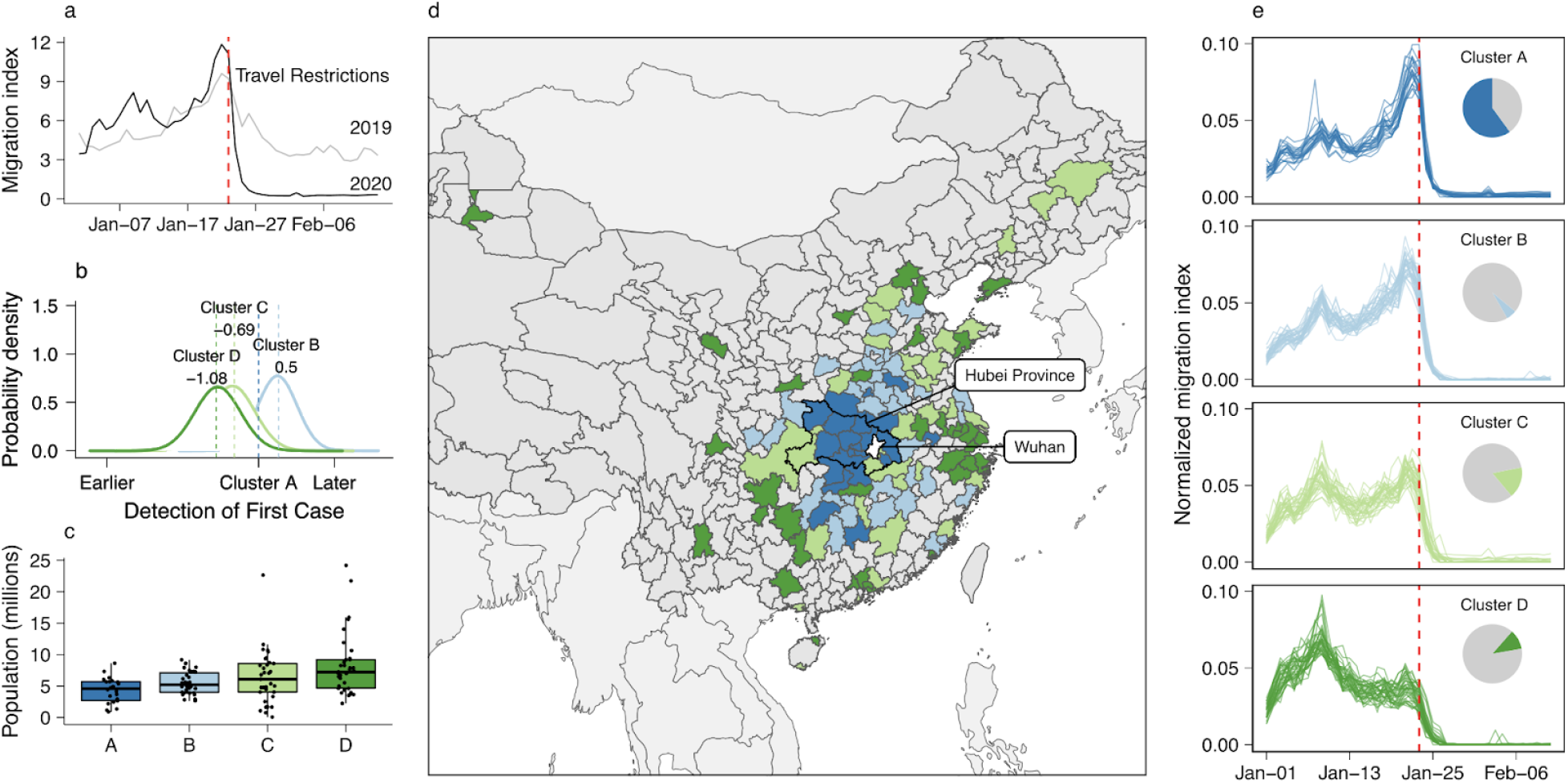
Travel patterns between Wuhan and its neighbors. The identified patterns of outbound travel from Wuhan: a), the daily outbound travel from Wuhan in 2019 and 2020; b), relative timing of first case detection stratified by clusters of similar trajectories, using Cluster A as the baseline. The distribution shows the mean effect size adjusted for surveillance intensity; c), distribution of resident population sizes of individual prefectures (points); d), map of prefectures and province-level cities showing the spatial distribution of trajectory clusters; e), outbound travel trends from Wuhan to the most connected prefectures in China, stratified by clusters of similar trajectories. The trajectories have been normalised by the total flow of each, to allow comparison of the profile. The pie charts show the total flux out of Wuhan prefecture by destinations in each cluster.

Using k-means clustering of the trajectories of outbound travel from Wuhan, we identified four general temporal clusters that captured the travel patterns from Wuhan to its neighbors (Figure 1e). Two of these clusters exhibited an increase in flow immediately before LNY (clusters A and B). Members of clusters A and B are geographically closer to Wuhan (Figure 1d), with fewer residents and overall lower population density (Figure 1e, and Table 3, Supplemental Table 1–3). Cluster C exhibited two peaks around 7 and 22 January 2020, respectively. Cluster D showed one peak in early-January 2020, with no peak immediately preceding the LNY holidays.The findings are not sensitive to the number of clusters, (Supplemental Figures 1–5).

The earliest detection of COVID-19 outside of Wuhan was 19th January 2020. Compared to Cluster A, Cluster B and C detected their first COVID-19 cases at approximately the same time (Fig 1d). Prefectures in cluster D confirmed their first cases 1.08 days earlier (Figure 1d). Cluster D includes large population centres; Beijing, Shanghai, Guangzhou, and Shenzhen. After the arrival of infected individuals from Wuhan, these highly connected cities could have contributed to the further spread of COVID-19 to places less directly connected to Wuhan. There were also a small number of prefectures that did not have any confirmed cases until 3 weeks after the *cordon sanitaire* in Wuhan.

We repeated the same analyses for other large cities in China, finding that despite the different numbers of clusters identified, the general patterns in movement flows observed in Wuhan were seen elsewhere in mainland China, with an early january peak travel, and another increase in travel volume preceding LNY (Supplemental Figures 6–10). The association between the population size of destinations and geographic distance, however, was less apparent. The early-January peak in Wuhan coincided with the beginning of winter break for university students in China, approximately one million of whom study in Wuhan^11^. Without information about the age composition of travellers at this time, we cannot provide a definite explanation of this observation.

By late March, over 90% of prefectures and province-level cities (further detail on administrative levels included in Methods) in mainland China had at least one confirmed case of COVID-19. Most prefectures confirmed their first COVID-19 cases between 23rd and 26th January 2020.

There is anecdotal evidence implying an association between the announcement of a *cordon sanitaire* on 23 January and temporarily increased outbound travel from Wuhan^12^. This relationship, if true, could have hindered the effectiveness of the *cordon sanitaire*. Focusing on the six-day period preceding the LNY, we compared the outbound travel patterns from Wuhan with the rest of mainland China using 2019 as the baseline. We used two variability metrics to investigate potential outbound travel surges: (1) a proportion-based matric (Methods, eq. 3) that captures the relative between-year difference; and (2) an anomaly-based metric (Methods, eq. 4) that captures the deviation observed in 2020 compared to 2019. We found that although there is evidence of an increase in outbound travel from Wuhan during this period, a similar increase was also observed in many other prefectures. Wuhan was ranked 46 (top 13%) and 88 (top 24%) of 305, by the two metrics for the change in flow (Supplemental Figure 11).

### Movement Patterns across China

To investigate the movement patterns across mainland China, we divided prefectures and province-level cities into four population quartiles (i.e. “Low” (2000 to 1.44 million residents), “Medium-low” (1.45 to 2.96 million residents), “Medium-high” (2.98 to 4.90 million residents), and “High” (4.92 to 24.20 million residents). We found that the trends of in- and outbound travel volume over time were relatively consistent across population quartiles (Supplemental Figure 12). The flow between all pairs of quartiles, measured in Baidu’s migration index, increased prior to LNY and dropped sharply after Wuhan’s *cordon sanitaire*, with an increase in within-quartile flow following 23rd January for all quartiles. However, the underlying composition of these in- and outbound travel flows differed substantially by population quartile (Figure 2).

**Figure 2.**
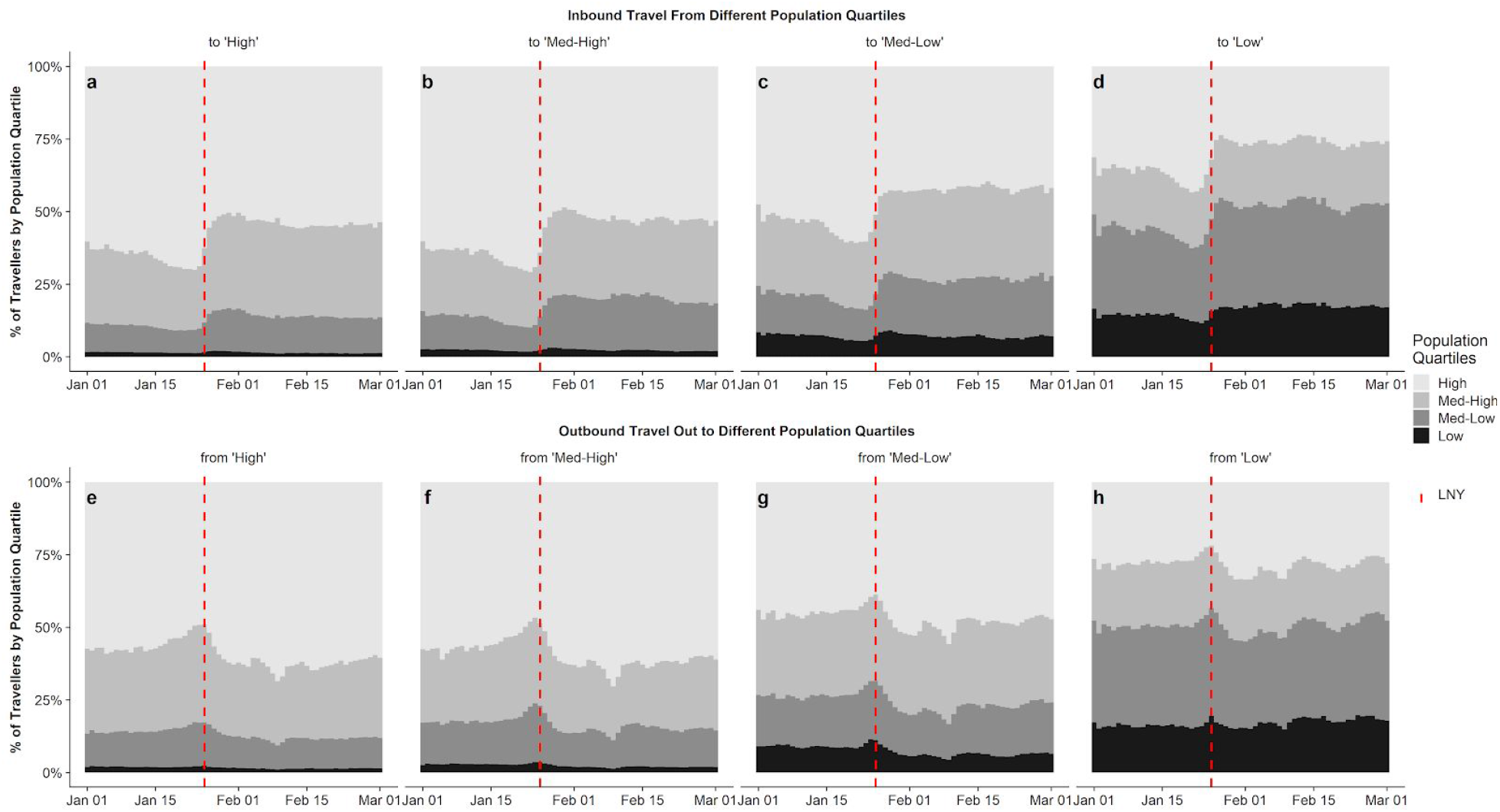
Contribution by prefectures of each population quartile to in- and outbound travel to different locations. Shading marks the population quartile with highest population quartiles in the lightest shade. Red dashed line shows the first day of LNY (25th January 2020).

Before LNY, all regions saw increased inbound travel from highly populated prefectures (Figure 2a-d). These changes were more marked in prefectures of lower population sizes. After LNY, the contribution to inbound travel by prefectures in the middle quartiles stabilised at higher levels compared to pre-LNY. As the volume of inbound travel recovered through February (Supplemental Figure 12), the relative proportion of travellers from the most populated quartiles remained low. For outbound travel, a higher proportion of travellers from the most populated prefectures travelled to the middle quartiles before LNY, and a higher proportion from medium-sized prefectures travelled to low-population prefectures (Figure 2e-h). Travel volumes and distance patterns in Beijing, Shanghai, Guangzhou began to return to normal more quickly than in Wuhan, and outbound travel generally recovered more after LNY (Supplemental Figure 13).

Analysis of origin or destination locations stratified by population sizes also revealed cascading effects: travellers from large prefectures more often travelled to other large or medium size prefectures; travellers from medium and small prefectures more often travelled between medium and small prefectures (Supplemental Figure 14). Therefore, medium sized locations could play a key role in limiting the spread of COVID-19 to prefectures with fewer residents.

### Healthcare availability at destination locations

The observed movement patterns have important public health implications: before LNY, the move away from population centres was also a migration away from settings with high access to healthcare, measured by the number of Grade II and III hospitals per 100,000 residents (Figure 3). Prefectures with higher access to healthcare had more outgoing than incoming travellers, and after LNY, travellers gradually returned to high healthcare access settings, but the overall geographic distribution of residents had not recovered to its pre-LNY conditions by 1st March 2020 (Figure 3a). This pattern was associated with COVID-19 related healthcare pressure, a measure of confirmed cases compared with healthcare availability (Figure 3b). From the week before LNY to two weeks after, locations with low access to healthcare experienced significantly higher pressure compared to locations with high access to healthcare. *Chunyun* not only increased the chance of infection along mobility networks, but also shifted healthcare pressure caused by COVID-19 to regions with low access to healthcare^13–15^.

**Figure 3.**
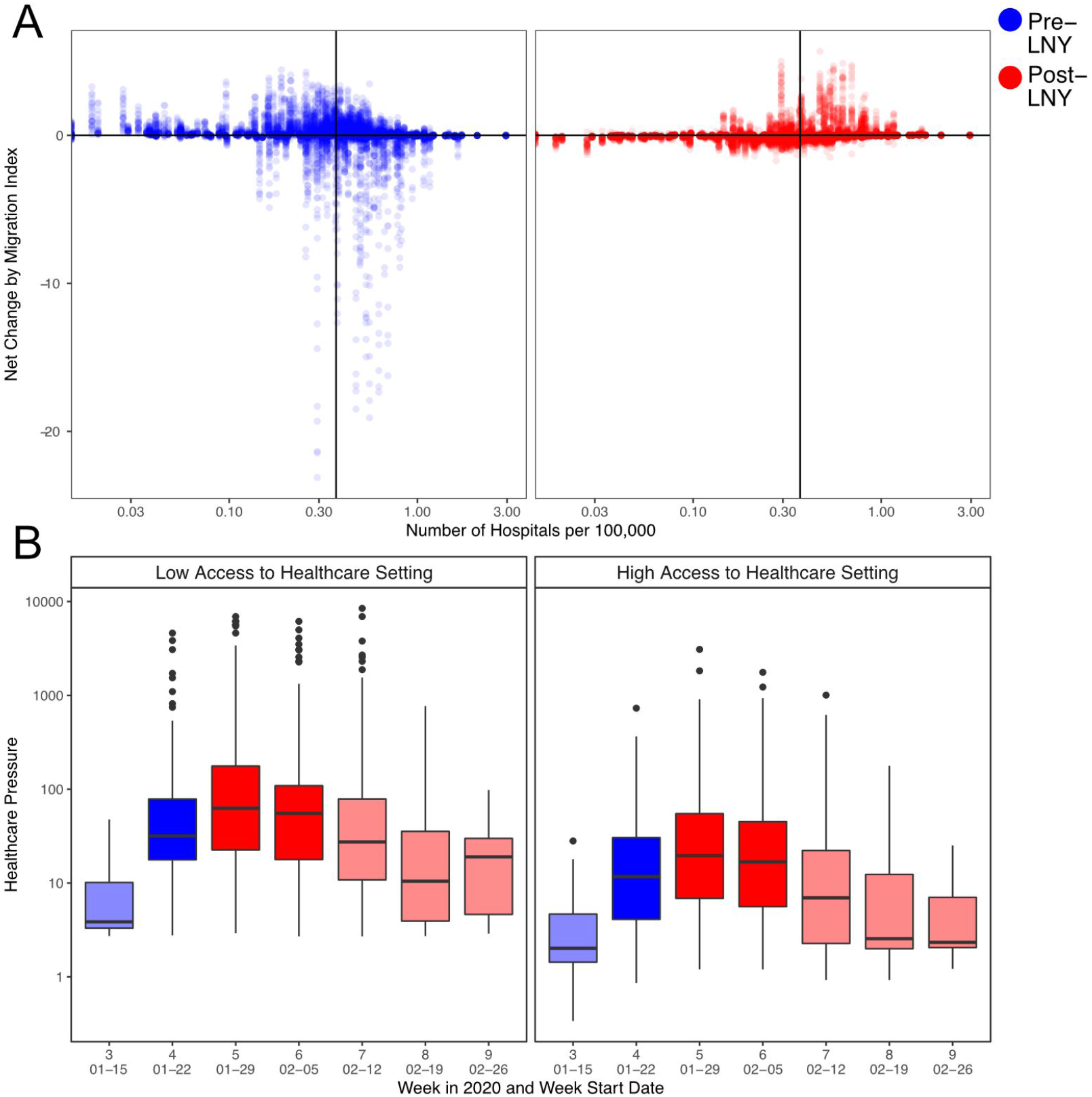
Human mobility, healthcare services availability, and COVID-19 related healthcare pressure. A), The changes in traveller volume before (blue) and after (red) LNY. Net change is defined as inbound migration index minus outbound migration index. Thus, a negative change indicates more travellers leave than arrive while a positive value indicates more travellers arrive than leave. Solid line indicates the median level of healthcare access. B), The changes in the healthcare pressure (log_10_ scale) related to COVID-19 each week in low and high healthcare access prefectures. Healthcare access is measured by the number of hospitals per 100,000 residents. Healthcare pressure is measured by confirmed COVID-19 cases divided by healthcare access. Darker shade represents weeks when low healthcare access settings experienced significantly higher pressure than high healthcare access settings; lighter shade represents when differences are not significant based on Mann-Whitney U test.

### Changes in overall travel network structure

We determined the community structure of the local travel network by calculating the daily modularity, Q, of the directed network^16^ from 1st January to 1st March 2020. This provides a holistic view of transport throughout the country, highlighting macroscopic changes in the network, e.g. rerouting behaviour or increased linkages between new prefectures, as the movement network adjusted to travel restrictions in Wuhan.

Preceding the implementation of travel restrictions, there was a stable pattern of communities connected to large cities, with significant flows between communities (Figure 4). Early January before LNY represents typical travel in China with flow between major population centres^17^. During this period, travel within China was generally structured into well-defined communities, with high Q (Figure 4, time point 1). Major cities had consistent, distinct communities over this time period, which remained fairly steady even as outflows began to increase from major cities for LNY (Figure 4, time point 2; see Supplement 6 for full time series).

**Figure 4.**
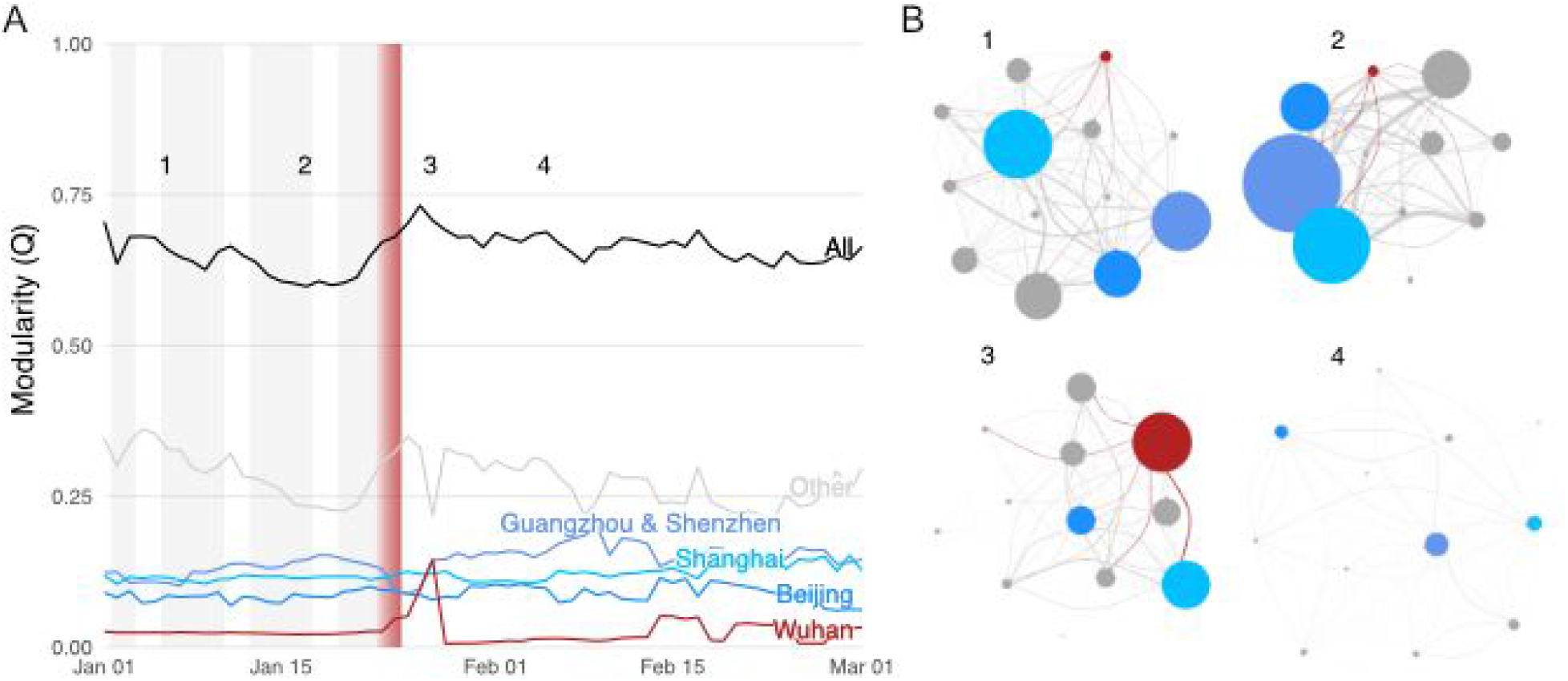
Community structure and modularity of movement network. The time series of total and sub-community modularity, and B) snapshots of the community networks on days before and after the *cordon sanitaire*. The communities for Wuhan (red) and several major cities (blues) are highlighted in A and B in both the community and edges between communities. The time series shows working week (grey bars; missing after nation-wide social distancing measures) as well as the initiation and enforcement of restrictions in Wuhan (red gradient) over 23rd to 24th January. The communities (circles) are sized according to within-community migration index, while their connections are sized according to their between community migration index.

Immediately following the implementation of travel restrictions, we identified a marked peak in modularity where the Q value for Wuhan City increased, indicating that it temporarily became more integrated into the travel network (Figure 4a, time point 3). This increase in modularity indicated relatively more connectivity between Wuhan and other communities, although there was decreased flow, so the actual number of travellers was much lower. This could also reflect the large movement of medical and other resources to Wuhan following the implementation of restrictions^18^.

Overall connectivity decreased across China after the *cordon sanitaire* in Wuhan (Figure 4, time point 4). This coincided with the implementation of disease control interventions in other prefectures, and a decrease in travel following LNY. Consistent with a country-wide policy of restricted movement, we did not find large rerouting or the increasing importance of other transport connections after the restrictions in Wuhan,. This is critical as countries attempt to determine the efficacy of large scale movement restrictions.

## Discussion

The *cordon sanitaire* in Wuhan was an intensive travel restriction that completely stopped all non-essential incoming and outgoing traffic. Previous studies have demonstrated that it may have had low effectiveness in preventing or delaying transmission to other regions of mainland China during the early phase of the COVID-19 pandemic^7,19^. There is however potential for infectious disease control and prevention, especially when timeliness and the necessary scope of restrictions can be achieved^20^. Travel restrictions will likely continue to be considered an important infectious disease intervention option against COVID-19 during the pandemic, and better understanding the mechanisms in play at different stages of travel restrictions is crucial to effective implementation.

We found a limited relationship between spatial proximity and epidemic spread where larger, distant populations detected their first COVID-19 cases earlier than smaller locations that are closer to Wuhan. Due to the highly connected modern mobility network, spatial proximity is not the only measure for “closeness” between two cities^21^. While planning for travel restrictions, either domestic or international, it may be worthwhile to consider other functional connectivity measures, such as human mobility studied here. Although outbreaks may appear to have single source location in the beginning, such as the case in Europe^22^, focussed travel restrictions around epicentres and their immediate geographic surroundings may lead to missed opportunities for epidemic control.

The timing of LNY and the initial stage of the COVID-19 epidemic makes it difficult to untangle regular holiday travel from travel in response to the outbreak or to impending travel restrictions. The increased outflow from Wuhan that we observed was not unique to the city, as similar patterns of outflow were observed in a large number of other prefectures, and so likely represents increased holiday travel. We therefore did not find evidence of an association between the announcement of the *cordon sanitaire* and the number of outbound travellers leaving Wuhan. Data from other countries not confounded by holiday travel (e.g., France^23^) may yield insights on public responses to travel restrictions. Additionally, although the overall number of travellers leaving Wuhan was not exceptionally high before LNY, the composition of travellers may have changed, such as a shift from business to family travel, which could contribute to the spread of COVID-19 and could have implications for healthcare demand in destination locations^24^. Finer resolution mobility data, including traveller characteristics such as age and occupation, could improve our understanding of the potential outbreak risk and the likely impacts of different interventions in the future.

Human mobility during *Chunyun* was marked by the general trend of people leaving large population centres for less populated locations, which was also migration away from locations with high access to healthcare. During the peak of the epidemics in mainland China, areas with low access to healthcare experienced significantly higher healthcare pressure related to COVID-19 compared to elsewhere. Temporarily mobilising resources such as medical personnel and equipment could aid epidemic control in places receiving a higher-than-normal number of travelers from places with potentially high COVID-19 prevalence, and thus could be evaluated as a potential public health intervention under similar circumstances^25^.

The structure of the overall transportation network in China did not demonstrate compensatory responses to the *cordon sanitaire*. There was a brief alteration of the network structure immediately following the restrictions, before the network settled quickly back into the same relatively stable communities that existed before the restrictions, albeit at markedly lower flow. This implies that the overall transportation network did not undergo structural reorganisation as a result of Wuhan’s *cordon sanitaire* and other regional travel restrictions. Short-term travel restrictions may therefore not incur lasting impacts on the mobility network, but assessing long-term impacts will require longer time-series analyses.

Mobility data from Baidu Huiyan has some limitations. For example, travel volumes were collected on an eight-hourly basis between each pair of prefectures and then aggregated to day- and prefecture-level, which does not allow analysis of trips longer than a day. In a country the size of China, such trips may be relatively frequent. Pairwise travel patterns before 1 January 2020 are not available, which makes it challenging to determine baseline travel patterns. Additionally, movement patterns from Baidu Huiyan reflect the movement of Baidu users, which may be a non-random subset of the general population in mainland China^26^.

This study analysed the human mobility patterns around China during different stages of the local COVID-19 epidemics, from early *Chunyun* to Wuhan’s *cordon sanitaire* and other travel restrictions. By the start of March 2020, regional inter-prefecture movements had started to recover. Many countries have now implemented similar travel restrictions to reduce disease transmission. Understanding the implications of travel patterns before, during, and following travel restrictions is valuable for informing public health interventions, surveillance, and healthcare demand planning globally.

## Methods

### Geographic Information

The geographic unit of analysis in this study is prefecture, which is administrative level 2 in mainland China, just below the province (level 1). There are currently more than 360 prefecture-level units in China. However, the four provincial level cities (Beijing, Tianjin, Shanghai, and Chongqing) are exceptions. They do not have a level 2 unit – level 1 directly manages level 3 administrative units (i.e., counties) in these locations^27^. In this study, we analysed these province-level cities with prefectures for spatial completeness.

### Mobility Data

The mobility data is publicly available through Baidu Huiyan^28^, a web service that supports government agencies and businesses with big-data spatio-temporal analytics. Estimates are based on over 120 billion location-based service (LBS) enquiries each day from over 1.1 billion mobile devices, while taking into consideration more than 1.5 billion points of interests (POI). We obtained two variables directly from Baidu Huiyan: overall migration index (specific to each prefecture) and percentage of travellers arriving in or leaving specific locations (specific to each pair of prefectures). Note that migration index is a relative measure of the magnitude of human mobility, scaled relative to the total volume of movement across the network.

We calculate the volume of human mobility between each pair of prefectures between 1st January 2020 and 1st March 2020 using the following equation:

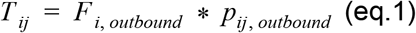

where *T_ij_* is the volume of mobility from location i to location *j*, *F* is the overall Baidu migration index with direction (*inbound* or *outbound*) at location *i*, and *p_ij_* is the proportion of all outbound travel that occurred between location *i* and location *j*. We further validated this measure by assuming that inbound and outbound were equal, as:

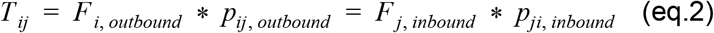

Note that *p_ij_* is only available from Baidu for the top 100 prefectures connected to *i*. In other words, *p_ij_* and *p_ji_* may not simultaneously exist, and so some values cannot be cross-validated. Using data from Baidu Huiyan, we created a 366*366 connectivity matrix for each day between 1 January 2020 and 1 Mar 2020 (61 days).

### Demographic and Healthcare System Data

The 2018 population sizes were retrieved from the China Statistics Yearbook^29^. The geographic boundaries of prefectures and province-level cities were obtained from the Institute of Geographic Sciences and Natural Resources Research (Chinese Academy of Sciences)^30^. The original source of daily confirmed incidence is the COVID-19 dashboard published by DXY.cn, which updates in near-real time based on government press releases^31^. Additionally, the package ‘nCoV2019’^32^ and ‘DXY-COVID-19-Crawler’^33^ have reduced the time required for data gathering and data cleaning. Records of first case arrivals were cross-checked with news articles also found on DXY.cn^31^. Information on the Grade II and III hospitals in China was retrieved from the National Health Commission^34^ and was then geo-referenced using the non-commercial Amap API^35^.

### Trajectory Analysis and Surge Evaluation

The patterns of movement out of Wuhan between 1st and 23rd January were analysed using trajectory cluster analysis of the magnitude-normalized trajectories of outflow over time. Outflow trajectories were selected by thresholding journeys with greater than an average flow index of 0.005 for the entire period. This threshold removed prefectures with negligible connectivity with the origin. In order to characterize the shape of the outflow trajectories, rather than the magnitude of certain outflows, outflow trajectories were normalised by dividing the flow for each day by the total migration between the 1st and 23rd January 2020 for each trajectory.

We classified the trajectories using k-means clustering with four clusters^36^. The number of clusters was chosen using a plot of average silhouette width against number of clusters, for between 4 and 12 clusters. The silhouette width decreased significantly at four clusters, and a similar number of trajectories were allocated to each cluster (Supplemental Figures 1–5). Furthermore, when using a greater number of clusters, we observed the same four overall trajectory patterns with smaller differences between trajectories defining each cluster. We also observed an increasingly large number of clusters containing a small number of trajectories. Plots of the trajectories clustered using 2, 3, 4, 5, and 6 clusters are included in the supplementary material. K-medioids, and Agglomerative Clustering were also explored as alternatives to K-means clustering. The different clustering methods did not result in substantial differences and identified similar patterns among outflow trajectories.

We also calculated the peak outflow from all prefectures 2 to 7 days before LNY (i.e., 0 to 5 days before the *cordon sanitaire*). We used two parameters to quantify the magnitude of change since 2019 at location *i*:

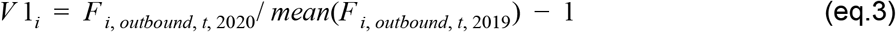

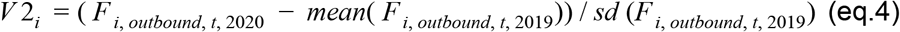

The variable *t* corresponds to 29 January – 3 February 2019 and 18 January – 23 January 2020.

The association between the arrival of the first case and travel clusters were explored via a linear model with two independent variables, travel cluster and population. Population sizes of prefectures were used as a proxy of surveillance intensity, i.e.we assumed that more public health surveillance was conducted in larger cities. The coefficients and the standard errors of travel clusters were then compared.

### Assessing Access to Healthcare

In this study, prefecture-level healthcare access was measured by the number of Grade II and III hospitals per 100,000 residents. In mainland China, Grade II and III hospitals have at least 100 hospital beds and are equipped with ventilators. They are more important compared to community hospitals and clinics for COVID-19 management due to high healthcare need in cases, and thus are a good indicator of healthcare access. Healthcare pressure was calculated by dividing weekly confirmed COVID-19 cases^31^ by the access to healthcare. The distributions of healthcare pressure were highly skewed, and we therefore used non-parametric one-tailed Mann-Whitney U test to compare the differences of healthcare pressure between low and high access to healthcare settings (n1 = 157, n2 = 153). This test was repeated for each week from week three to nine, and the corresponding *p* –values are 8.75e-1, 1.98e-7, 1.21e-8, 3.67e-5, 5.63e-2, 4.86e-1, and 9.26e-1.

### Network Analysis

Using the weighted movement flows between locations, we calculated community structure in the network using the Leiden algorithm^16^. The Leiden algorithm maximizes the modularity, *Q*, on directed, weighted, time sliced networks with an inter-slice weighting of 10^−5^, which is the order of magnitude of minimum intra-slice weight across all times^37^. Modularity is a metric of within-community vs between-community connectivity, and the algorithm detects communities by optimising the within vs between, thereby assigning nodes to communities. Using the community structure from this algorithm, we identified the relative contributions to modularity, *Q*, of 4 key communities: the community containing Wuhan prefecture, and then the communities of four other major cities in China: Beijing, Shanghai, Guangzhou, and Shenzhen. The latter two were always assigned to the same community and are marked together in Figure 4. We presented 4 snapshots of communities in the travel network, but all are shown in Supplemental Figure 15, and the spatial locations of those networks in Supplemental Figure 16.

### Sensitivity Analyses

We repeated clustering of temporal traveller flow trajectories to validate the method for assessing travel flux out of Wuhan between January 1st and January 23rd. Employing the same method of thresholding prefectures with little connectivity, the trajectories of travellers to individual destination locations over time were normalised by dividing by the total flow along each route in the period. These normalized trajectories were then clustered using the same k-means clustering procedure discussed above. The number of clusters was determined using a silhouette plot in order to isolate the dominant temporal patterns of traveller movement to individual destinations.

## Data Availability

All data used were publicly available and can be found at the references. Code is publicly available in a Github repository.

https://github.com/yangclaraliu/pandemic_travel_china

## Author Contributions

HG, YL, RME, conceived the methods in the study. HG, YL, CIJ, and CABP implemented the analysis and generated the figures with input from RME & CG. All authors interpreted the findings and prepared the manuscript. All authors reviewed the manuscript and approved the final version for submission.

## Funding

The authors acknowledge funding as follows: HG: funding from the Department of Health and Social Care using UK Aid funding and managed by the NIHR under grant (ITCRZ 03010). YL: Bill and Melinda Gates Foundation (INV-003174), NIHR (16/137/109), European Commission (101003688); CABP: BMGF (OPP1184344) & DFID / Wellcome (ref. 221303/Z/20/Z); CIJ: Global Challenges Research Fund (GCRF) project ‘RECAP’ managed through RCUK and ESRC (ES/P010873/1); BQ This research was partly funded by the National Institute for Health Research (NIHR) (16/137/109) using UK Aid from the UK Government to support global health research; CD: NIHR (16/137/109); and RME: HDR UK (grant: MR/S003975/1), MRC (grant: MC_PC 19065).

The views expressed in this publication are those of the author(s) and not necessarily those of the NIHR or the UK Department of Health and Social Care.

## The working group acknowledges the following funding

QJL: Medical Research Council London Intercollegiate Doctoral Training Program studentship (grant no. MR/N013638/1), AG: Global Challenges Research Fund (GCRF) for the project “RECAP” managed through RCUK and ESRC (ES/P010873/1), MA: Bill and Melinda Gates Foundation (OPP1191821), RL: Royal Society Dorothy Hodgkin Fellowship, KO’R: Bill and Melinda Gates Foundation (OPP1191821), MQ: European Research Council Starting Grant (Action Number #757699), JH: Wellcome Trust (grant: 210758/Z/18/Z), GMK: UK Medical Research Council (grant: MR/P014658/1), TJ: RCUK/ESRC (grant: ES/P010873/1); UK PH RST; NIHR HPRU Modelling Methodology, PK: This research was partly funded by the Royal Society under award RP\EA\180004, European Commission (101003688), Bill & Melinda Gates Foundation (INV-003174), SRP: Bill and Melinda Gates Foundation (OPP1180644), EMR: Medical Research Council London Intercollegiate Doctoral Training Program studentship (grant MR/N013638/1), SFlasche: Wellcome Trust (grant: 208812/Z/17/Z), AJK: Wellcome Trust (grant: 206250/Z/17/Z), SA: Wellcome Trust (grant: 210758/Z/18/Z), FYS: NIHR EPIC grant (16/137/109), AE: The Nakajima Foundation; The Alan Turing Institute, GM: NTD Modelling Consortium by the Bill and Melinda Gates Foundation (OPP1184344), JDM: Wellcome Trust (grant: 210758/Z/18/Z), SRM: Wellcome Trust (grant: 210758/Z/18/Z), NIB: Wellcome Trust (grant: 210758/Z/18/Z), WJE: European Commission (101003688), NGD: (NIHR: Health Protection Research Unit for Modelling Methodology HPRU-2012–10096), KP: Gates (INV-003174), European Commission (101003688), CJVA: European Research Council Starting Grant (Action Number #757688), ESN: Gates (OPP1183986), RMGJH: European Research Council Starting Grant (Action Number #757699), JCE: European Research Council Starting Grant (Action Number #757699), KvZ: KvZ, is supported by the UK Government Department for International Development (DFID)/Wellcome Trust Epidemic Preparedness Coronavirus research programme (ref. 221303/Z/20/Z), and Elrha’s Research for Health in Humanitarian Crises (R2HC) Programme, which aims to improve health outcomes by strengthening the evidence base for public health interventions in humanitarian crises. The R2HC programme is funded by the UK Government (DFID), the Wellcome Trust, and the UK National Institute for Health Research (NIHR), KEA: European Research Council Starting Grant (Action Number #757688), AR: NIHR (grant: PR-OD-1017–20002), SFunk: Wellcome Trust (grant: 210758/Z/18/Z), MJ: Gates (INV-003174), NIHR (16/137/109), European Commission (101003688), SC: Wellcome Trust (grant: 208812/Z/17/Z), TWR: Wellcome Trust (grant: 206250/Z/17/Z).

## Competing interests

Authors declare no competing interests.

## Data availability

All data used were publicly available and can be found at the references.

## Code availability

Code is publicly available in a Github repository, https://github.com/yangclaraliu/pandemic_travel_china.

## Notes

### Competing Interest Statement

The authors have declared no competing interest.

## References

1. Wang, X., Liu, C., Mao, W., Hu, Z. & Gu, L. Tracing The Largest Seasonal Migration on Earth. arXiv [physics.soc-ph] (2014).

2. Xinhua News Agency. This year’s Spring Festival passenger shipments will reach 2.99 billion. Gov.cn http://www.gov.cn/xinwen/2019-01/07/content_5355661.htm (2019).

3. Daily, B. Y. Go home early on the first day of the Spring Festival. Xinhuanet http://www.xinhuanet.com/2020-01/11/c_1125448045.htm (2020).

4. Shi, P., Keskinocak, P., Swann, J. L. & Lee, B. Y. The impact of mass gatherings and holiday traveling on the course of an influenza pandemic: a computational model. BMC Public Health 10, 778 (2010).

5. Diary of Pneumonia | January 23: Wuhan is fully ‘closing the city’ to fully contain the epidemic. Caixin http://www.caixin.com/2020-01-23/101507939.html (2020).

6. Lai, S. et al. Effect of non-pharmaceutical interventions for containing the COVID-19 outbreak in China. Infectious Diseases (except HIV/AIDS) (2020) doi:10.1101/2020.03.03.20029843.

7. Kraemer, M. U. G. et al. The effect of human mobility and control measures on the COVID-19 epidemic in China. Science (2020) doi:10.1126/science.abb4218.

8. Tian, H. et al. An investigation of transmission control measures during the first 50 days of the COVID-19 epidemic in China. Science (2020) doi:10.1126/science.abb6105.

9. Pepe, E. et al. COVID-19 outbreak response: a first assessment of mobility changes in Italy following national lockdown. medRxiv 2020.03.22.20039933 (2020).

10. Baidu. Baidu Huiyan Map. Baidu https://huiyan.baidu.com/.

11. Wuhan University. Notice about winter vacation in 2020. https://www.whu.edu.cn/info/1118/13775.htm https://www.whu.edu.cn/info/1118/13775.htm (2020).

12. 5 million left Wuhan before lockdown, 1,000 new virus cases expected. South China Morning Post https://www.scmp.com/news/china/society/article/3047720/chinese-premier-li-keqiang-head-coronavirus-crisis-team-outbreak (2020).

13. Bengtsson, L., Lu, X., Thorson, A., Garfield, R. & von Schreeb, J. Improved response to disasters and outbreaks by tracking population movements with mobile phone network data: a post-earthquake geospatial study in Haiti. PLoS Med. 8, e1001083 (2011).

14. Lu, X., Bengtsson, L. & Holme, P. Predictability of population displacement after the 2010 Haiti earthquake. Proc. Natl. Acad. Sci. U. S. A. 109, 11576–11581 (2012).

15. Five Findings from a New Phone Survey in Senegal. Center For Global Development https://www.cgdev.org/blog/five-findings-new-phone-survey-senegal.

16. Traag, V. A., Waltman, L. & van Eck, N. J. From Louvain to Leiden: guaranteeing well-connected communities. Sci. Rep. 9, 1–12 (2019).

17. Lai, S. et al. Assessing spread risk of Wuhan novel coronavirus within and beyond China, January-April 2020: a travel network-based modelling study. Epidemiology (2020) doi:10.1101/2020.02.04.20020479.

18. People. 346 medical teams of 42,000 people arrived in Hubei to fight the epidemic. People’s Daily Overseas Edition http://paper.people.com.cn/rmrbhwb/html/2020-03/09/content_1975139.htm (2020).

19. Zhanwei Du et al. Risk for Transportation of Coronavirus Disease from Wuhan to Other Cities in China. Emerging Infectious Disease journal 26, 1049 (2020).

20. Bajardi, P. et al. Human mobility networks, travel restrictions, and the global spread of 2009 H1N1 pandemic. PLoS One 6, e16591 (2011).

21. Iannelli, F., Koher, A., Brockmann, D., Hövel, P. & Sokolov, I. M. Effective distances for epidemics spreading on complex networks. Phys Rev E 95, 012313 (2017).

22. Grosso, L. In Italy’s coronavirus epicenter, life is on hold. POLITICO https://www.politico.eu/article/italy-coronavirus-covid19-lombardy-lodi/ (2020).

23. Tidman, Z. More than one million people fled Paris as coronavirus lockdown began. The Independent (2020).

24. Ewing, A., Lee, E. C., Viboud, C. & Bansal, S. Contact, Travel, and Transmission: The Impact of Winter Holidays on Influenza Dynamics in the United States. J. Infect. Dis. 215, 732–739 (2017).

25. WHO. Strengthening the health system response to COVID-19 Recommendations for the WHO European Region Policy brief. http://www.euro.who.int/__data/assets/pdf_file/0003/436350/strengthening-health-system-response-COVID-19.pdf (2020).

26. Chyxx. Analysis of the development of mobile phone Baidu in 2017. Chyxx https://www.chyxx.com/industry/201804/625950.html (2018).

27. Ministry of Civil Affairs of the People’s Republic of China. 2020 administrative division cod. Ministry of Civil Affairs of the People’s Republic of China http://www.mca.gov.cn/article/sj/xzqh/2020/.

28. Baidu. Baidu Migration. https://qianxi.baidu.com/ https://qianxi.baidu.com/.

29. China Statistical Yearbook 2018. http://www.stats.gov.cn/tjsj/ndsj/2018/indexeh.htm.

30. Chinese Academy of Sciences. Institute of Geographic Resources. Institute of Geographic Sciences and Natural Resources http://www.igsnrr.ac.cn/.

31. DXY. COVID-19 Global Pandemic Real-time report. ncov.dxy https://ncov.dxy.cn/ncovh5/view/pneumonia.

32. Yu, G. nCov2019. (Github).

33. Lin, I. DXY-COVID-19-Crawler. (Github).

34. China, C. D. C. Public Health Science Data Center. China CDC data center http://www.phsciencedata.cn/Share/.

35. Alibaba. IBS Amap API. IBS AMAP https://lbs.amap.com/.

36. Bishop Christopher, M. Pattern recognition and machine learning. Information science and statisticsNew York: Springer (2006).

37. Mucha, P. J., Richardson, T., Macon, K., Porter, M. A. & Onnela, J.-P. Community structure in time-dependent, multiscale, and multiplex networks. Science 328, 876–878 (2010).

